# Neutrophil-mediated Oxidative Stress and Albumin Structural Damage Predict COVID-19-associated Mortality

**DOI:** 10.1101/2021.04.01.21254767

**Authors:** Mohamed A. Badawy, Basma A. Yasseen, Riem M. El-Messiery, Engy A. Abdel-Rahman, Aya A. Elkhodiry, Azza G. Kamel, Asmaa M. Shedra, Rehab Hamdy, Mona Zidan, Diaa Al-Raawi, Mahmoud Hammad, Nahla Elsharkawy, Mohamed El Ansary, Ahmed Al-Halfawy, Alaa Elhadad, Ashraf Hatem, Sherif Abouelnaga, Laura L. Dugan, Sameh S. Ali

**Affiliations:** Research Department, Children’s Cancer Hospital Egypt 57357, Cairo, Egypt; Infectious Disease Unit, Internal Medicine Department, Faculty of Medicine, Cairo University, Cairo, Egypt; Pharmacology Department, Faculty of Medicine, Assuit University, Assuit, Egypt; Pediatric Oncology Department, National Cancer Institute, Cairo University and Children’s Cancer Hospital 57357, Cairo, Egypt; Clinical pathology department, National Cancer Institute, Cairo University and Children’s Cancer Hospital 57357, Cairo, Egypt; Department of Intensive Care, Faculty of Medicine, Cairo University, Cairo, Egypt; Department of Pulmonary Medicine, Faculty of Medicine, Cairo University, Cairo, Egypt; Department of Chest Diseases, Faculty of Medicine, Cairo University, Cairo, Egypt; Division of Geriatric Medicine, Department of Medicine, Vanderbilt University Medical Center; and VA Tennessee Valley Geriatric Research, Education and Clinical Center (GRECC), Nashville, TN, United States

**Keywords:** Critically ill COVID-19 patients, COVID-19 Mortality, Neutrophils, Oxidative Stress, Human Serum Albumin Damage, Spin Labels, Electron Paramagnetic Resonance

## Abstract

Human serum albumin (HSA) is the frontline antioxidant protein in blood with established anti-inflammatory and anticoagulation functions. Here we report that COVID-19-induced oxidative stress inflicts structural damages to HSA and is linked with mortality outcome in critically ill patients. We recruited 25 patients who were followed up for a median of 12.5 days (1-35 days), among them 14 had died. Analyzing blood samples from patients and healthy individuals (n=10), we provide evidence that neutrophils are major sources of oxidative stress in blood and that hydrogen peroxide is highly accumulated in plasmas of non-survivors. We then analyzed electron paramagnetic resonance (EPR) spectra of spin labelled fatty acids (SLFA) bound with HSA in whole blood of control, survivor, and non-survivor subjects (n=10-11). Non-survivors’ HSA showed dramatically reduced protein packing order parameter, faster SLFA correlational rotational time, and smaller S/W ratio (strong-binding/weak-binding sites within HSA), all reflecting remarkably fluid protein microenvironments. Stratified at the means, Kaplan–Meier survival analysis indicated that lower values of S/W ratio and accumulated H_2_O_2_ in plasma significantly predicted in-hospital mortality (S/W<0.16, 80% (9/12) vs. S/W>0.16, 20% (2/10), *p*=0.008; plasma [H_2_O_2_]>7.1 μM, 83.3% (5/6) vs. 16.7% (1/6), *p*=0.049). When we combined these two parameters as the ratio ((S/W)/[H_2_O_2_]) to derive a risk score, the resultant risk score lower than the mean (< 0.0253) predicted mortality with 100% accuracy (100% (6/6) vs. 0% (0/6), logrank χ^2^ = 12.01, p = 5×10^−4^). The derived parameters may provide a surrogate marker to assess new candidates for COVID-19 treatments targeting HSA replacements.

## Introduction

COVID-19 pandemic continues as a global health crisis while the underlying SARS-CoV-2 virus defies all attempted treatment strategies. While writing this report there have been more than 135 million confirmed cases including around 3 million deaths worldwide according to the World Health Organization Coronavirus Disease Dashboard (https://covid19.who.int/). Although 50% of cases are reported to be in the 25-64 age group, the percentage of deaths increases dramatically with age, and approximately 75% of deaths are in those aged 65 years and above ([COVID-19 Hospitalization and Death by Age | CDC](https://www.cdc.gov/coronavirus/2019-ncov/covid-data/investigations-discovery/hospitalization-death-by-age.html)). People in the age groups 30-39 years, 40-49 years, and 50-64 years are 4, 10, and 30 times more likely to die from COVID-19 complications compared to 18-29 years’ age group. Nevertheless, molecular and cellular factors contributing to mortality outcome in a homogeneous cohort of patients are not yet clear. Lack of diagnostic markers that predict mortality in COVID-19 patients impedes current efforts to siege the pandemic. It is thus critical to identify prognostic tests that can assess the risk of death in critically ill patients to guide clinical protocols and prioritize interventions. Furthermore, mechanistic clues for determining the underlying molecular factors contributing to the hypercoagulability, inflammation, and cytokine storm have been so far illusive. It is therefore imperative to intensify efforts focusing on understanding the molecular pathophysiology of COVID-19 infection and to identify prognostic markers to guide and prioritize clinical decisions.

Human serum albumin (HSA) is the most abundant constituent of soluble proteins in the circulatory system. HSA has been suggested and used as a diagnostic and prognostic marker of numerous diseases and conditions including ischemia, rheumatoid arthritis, cancer, septic shock among many others. In addition to its numerous physiological and pharmacological functions including the maintenance of blood/tissue osmotic balance (1), blood pH, metal cation transport and homeostasis (2, 3), nutrients and drug shuttling (4, 5), toxin neutralization (6, 7), HSA is suggested to be a major circulating antioxidant (8, 9). HSA can remarkably bind with a diverse array of drugs and toxins thus controlling their bioavailability and pharmacologic effects (10). It has been previously shown that more than 70% of the free radical-trapping capacity of serum was due to HSA (reviewed in (11)). Importantly, several reports indicated that inflammation enhances vascular permeability of various tissues to HSA apparently to confer antioxidant beneficial effects against reactive species released by activated neutrophils (12-14). Although currently without direct experimental evidence, neutrophilia-mediated oxidative stress was implicated in the COVID-19 pathology and speculated to exacerbate the inflammatory immune response eventually causing multi organ failure and death (15). We hypothesized that COVID-19-mediated oxidative stress may be differentially reflected in HSA’s structure and functions and employed electron paramagnetic resonance (EPR) spin labeling spectroscopy to explore HSA’s structural changes in correlation with severity and mortality of critically ill COVID-19 patients.

Spin labeled long-chain fatty acids (SLFA) are established probes to explore structural and functional changes in albumin by EPR spectroscopy (16, 17). This approach relies on the well-studied ability of albumin to strongly and exclusively bind with fatty acids in blood. Albumin has at least seven different specific binding sites for long chain fatty acids located in different domains within the protein (18-20). Effectively, structural and functional changes in HSA may be assessed through the detection of parallel changes in mobility and binding affinity of SLFAs, in addition to the distribution of the spin labels on the albumin molecule (17). EPR spectra of spin labels bound to different domains of the protein provide information on the local fatty acids/protein interactions, which may probe changes in the overall structure of the protein under unfolding or damaging conditions; Fig. 1A, (18). Here we compare changes that occur to the mobility, binding affinity, and distribution of the HSA-bound SLFA in whole blood and plasma from COVID-19 patients in critical care unit relative to those observed in normal healthy individuals.

**Fig. 1.**
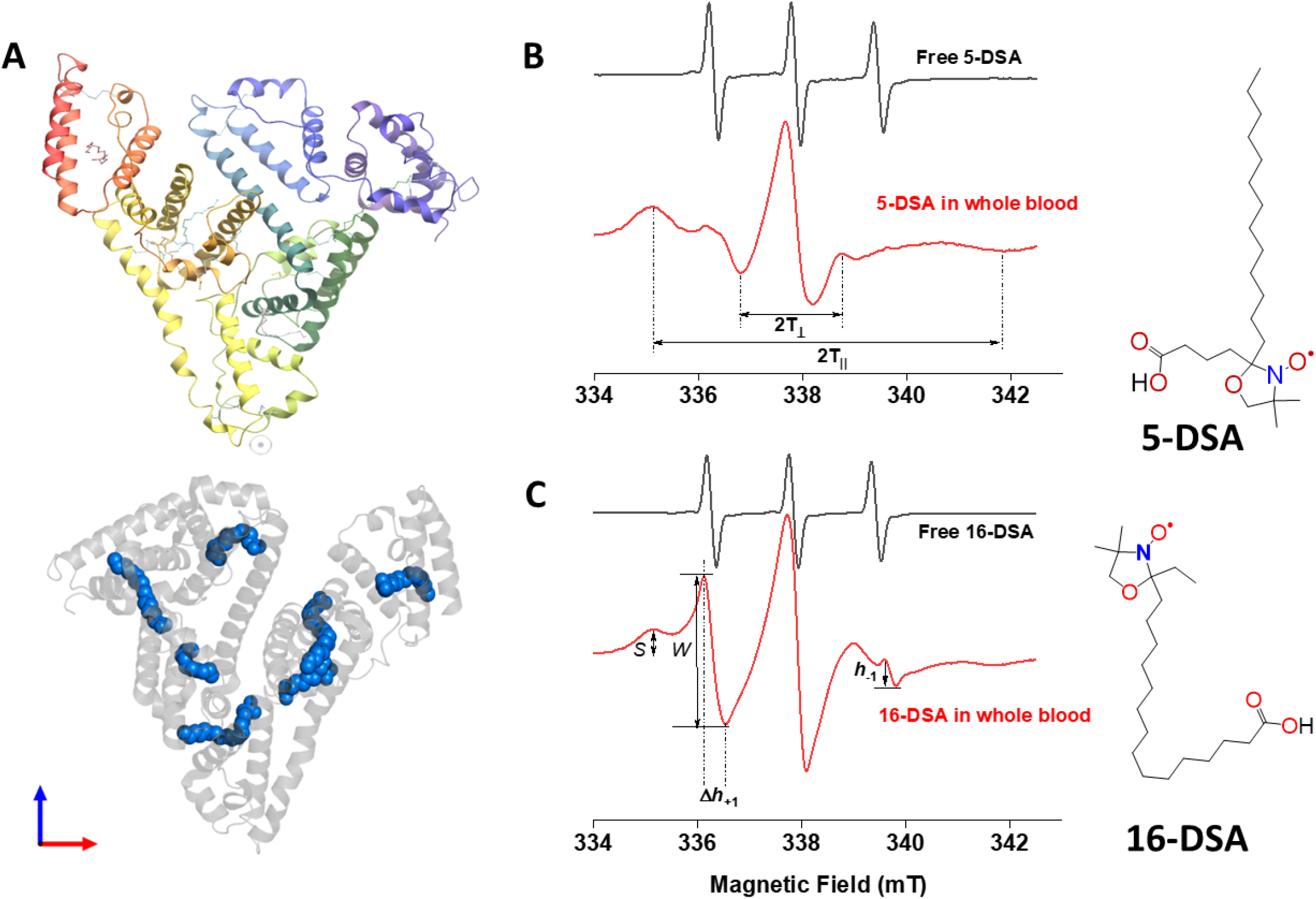
A) HSA crystal structure containing 7 copies of stearic acid. B) Representative EPR spectra of free and HSA-bound 5-DSA (B) and 16-DSA (C) in whole blood from the same COVID-9 recovered patient. Chemical structures of the two spin-labeled fatty acids are given on the right side of the figure.

## Methods

### Study design and participants

The present study aims to analyze HSA protein configuration statuses in the most severe cases of COVID-19 in comparison with control subjects. This is a prospective observational cohort study of patients with confirmed RT-PCR positive COVID-19. Nasopharyngeal swab RT-PCR results and Lung CT scans were combined to classify severe symptomatic COVID-19 cases. All patients were recruited from Kasr Alainy Cairo University Hospital/ ICU-facility at the Internal Medicine Quarantine Hospital. Supportive therapy including supplemental oxygen and symptomatic treatment were administered as required. Patients with moderate to severe hypoxia (defined as requiring fraction of inspired oxygen [FiO2] ≥40%) were transferred to the intensive care for further management including invasive mechanical ventilation when necessary. Patients recruited in the current study were divided into two arms based on future mortality outcome: Those survived past 10 days following blood samples collection (Sev-R) and those who died within 10 days of samples collection (Sev-D).

No power analysis was done. Sample size was based on sample availability and collection continued until a total of N=25 has been reached for COVID-19 patients diagnosed as severely infected with SARS-CoV-2 and admitted to the ICU during the period from 13 October 2020 to 25 January 2021. Within the follow-up time 14 patients had died. However, in a few cases and due to leukopenia or blood samples insufficiency to run every experiment on each subject, a number of 6-11 were analyzed and reported per condition. Nevertheless, all available samples were investigated by blinded operators and were all included in the final analysis. Demographic and clinical data of the studied 25 COVID-19 confirmed positive cases were categorized according to mortality and used for correlative analyses (Table 1). No randomization was done and clinical data were made available after EPR data collection and analysis have been performed. Patients who survived 10-days post sample collections were considered survivors (Sev-R) while those died within this period were placed in the Sev-D group.

**Table 1.** Demographic and clinical characteristics of the studied subjects.

|  | Sev-R | Sev-D | Tukey 95%-CI | *p* |
| --- | --- | --- | --- | --- |
| **n** | 11 | 14 |  |  |
| **Age (mean ± SD)** | 62.6 ± 9.7 | 69.8 ± 7.0 | 0.33-14.11 | 0.04 |
| **Male** | 54.54% | 57.14% |  | 0.896† |
| **sO<sub>2</sub> (mean ± SD)** | 78.57 ± 13.6 | 73.71 ± 17.07 | -20.42–10.7 | 0.52 |
| **Hypertension** | 27.27% | 42.85% |  | 0.65† |
| **Diabetes** | 18.18% | 50% |  | 0.09† |
| **Cardiovascular disease** | 0% | 14.28% |  | 0.2† |
| **Cancer** | 0% | 7.14% |  | 0.36† |
| **Bronchial asthma** | 0% | 7.14% |  | 0.36† |
| **ACE inhibitors** | 0% | 7.14% |  | 0.36† |
| **ARBs** | 9.09% | 7.14% |  | 0.85† |
| **calcium channel blocker** | 9.09% | 7.14% |  | 0.85† |
| **Beta Blockers** | 0% | 7.14% |  | 0.36† |
| **Diuretics** | 0% | 7.14% |  | 0.36† |
| **Sulphonylurea** | 9.09% | 21.42 |  | 0.4† |
| **Biguanides** | 0% | 7.14% |  | 0.36† |
| **Insulin** | 18.18% | 7.14% |  | 0.39† |
| **Anticoagulant** | 27.27% | 35.71% |  | 0.65† |
| **Steroids** | 45.45% | 57.14% |  | 0.56† |
| **Hydroxychloro-quine** | 18.18% | 7.14% |  | 0.4† |
| **IL-6 receptor antibody** | 9.09% | 35.71% |  | 0.12† |
| **Proton-pump inhibitor** | 9.09% | 14.28% |  | 0.69† |
| **Azithromycin** | 9.09% | 28.57% |  | 0.22† |
| **Cephalosporin** | 18.18% | 14.28% |  | 0.79† |
| **Carbapenem** | 9.09% | 42.85% |  | 0.06† |
| **Oxazolidinone** | 27.27% | 35.71% |  | 0.65† |
| **Fluoro-quinolone** | 36.36% | 35.71% |  | 0.97† |
| **Nitrofurantoin** | 9.09% | 0% |  | 0.25† |
| **Remdesivir** | 0% | 35.71% |  | 0.026† |
| **Ivermectin** | 0% | 21.42% |  | 0.10† |

sO<sub>2</sub>, blood oxygen saturation level, ACE, angiotensin converting enzyme; ARBs, angiotensin receptor blockers; IL-6, interleukin-6. † *p* values obtained through Pearson's $\chi^2$ test.

Written informed consents were obtained from participants in accordance with the principles of the Declaration of Helsinki. For COVID-19 and control blood/plasma collection, Children’s Cancer Hospital’s Institutional Review Board (IRB) has evaluated the study design and protocol, IRB number 31-2020 issued on July 6, 2020.

### Electron paramagnetic resonance (EPR) measurements

We measured EPR spectra at 37 °C using a Benchtop Magnettech MiniScope MS5000 spectrometer (now Bruker Biospin, Berlin) equipped with Biotemperature control and computerized data acquisition and analysis capabilities. Typical instrumental parameters during these measurements were: microwave frequency 9.47 GHz, microwave power 10 mW, modulation frequency 100 kHz, modulation amplitude 0.2 mT, magnetic field range 332-342 mT. Each spectrum was the average of 5 scans with scan time of 60 s. Spin-labelled stearic acids (Fig. 1): 2-(3-carboxypropyl)-4,4-dimethyl-2-tridecyl-3-oxazolidinyloxy (5-doxyl-stearic acid, 5-DSA) and 2-(14-carboxytetradecyl)-2-ethyl-4,4-dimethyl-3-oxazolidinyloxy (16-doxyl-stearic acid, 16-DSA) were purchased from Aldrich Chemical Co. (Milwaukee, U.S.A.). Spin labeled fatty acid (SLFA)-serum albumin complexes were formed as recommended (17) in 0.01 M phosphate buffer (pH 7.4). Conditions were optimized to avoid aggregation of free SLFA and to minimize contributions from albumin-unbound SLFA. Signals were stable over the measurement time and at least up to 1 h afterward as observed by following time evolution of the recorded EPR spectra.

### Analysis of biophysical EPR parameters

Fluidity measurement was carried out as we reported (21) and explained (22) previously. Nitroxyl radicals SLFA probes were used to determine local fluidity near the protein/aqueous interface (5-DSA) or the hydrophobic protein cores (16-DSA) of HSA. Whole blood or collected plasma from all subjects were labelled with 5-DSA or 16-DSA. Ethanolic solutions (0.02 M) of the spin labels were added to the blood/plasma in the ratio 1:100 and the cells were measured at 37 °C.

From the spectrum of 5-DSA an order parameter (*S*) was derived by measuring the outer and inner hyperfine splitting 2*T*_‖_ and 2*T*_⊥_ as defined in Fig. 1B (23, 24) using the formula:

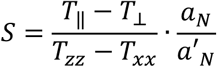

where

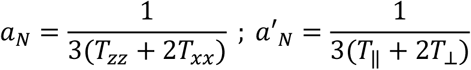

*T*_xx_ and *T*_zz_ are principal values of hyperfine tensor T taken as 0.61 mT and 3.24 mT; respectively (24, 25). To estimate fluidity of the inner lipophilic protein compartment, we used 16-DSA, i.e. a spin probe containing the nitroxide group attached on C16 that is located on the opposite terminal relative to the charged carboxyl fatty acid terminus. Rotational correlation time (*τ*_c_), which is taken as a measure of microenvironment fluidity in the fatty acid carrier sites in the HSA protein core was calculated as (26):

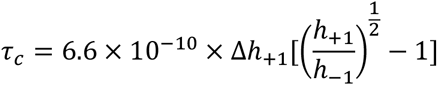

Where the parameters Δ*h*_+1_, *h*_+1_, and *h*_-1_ are determined from EPR spectra as shown in Fig. 1C. 16-DSA spectra (Fig. 1C) were also analyzed to obtain the ratio between weakly (W) and strongly (S) bound subpopulations of the spin label in sterically hollow versus relatively jammed protein microenvironments (22, 24).

### Phenotyping of peripheral blood by flow cytometry

Citrate-anticoagulated whole peripheral blood was incubated with RBCs lysis buffer for 15 min. Lysed blood was then centrifuged at 500 X-g for 5 min. Cells were washed twice with phosphate buffered saline (PBS) and then resuspended in PBS. Distribution status of platelets, neutrophils, monocytes and lymphocytes was measured in whole blood samples by 13-color flow cytometry as described using CytoFLEX system (Beckman Coulter Life Sciences CytoFLEX benchtop flow cytometer). Suspended cells were incubated with combinations of anti-human monoclonal antibodies for subset identification as follows: CD-42b-PE (Beckman Coulter life sciences, IM1417U) for platelets, CD14-PC7 (Beckman Coulter life sciences, A22331) for monocytes, CD66b-APC-Alexa Fluor 750 for neutrophils (Beckman Coulter life sciences, B08756) and CD3-ECD (Beckman Coulter life sciences, IM2705U) for lymphocytes cells were incubated for 30 minutes in the dark at room temperature. At the end of the incubation period, cells were washed with PBS and resuspended in 300 μl PBS. Samples were then analyzed by flow cytometry for gating platelet specific CD42b-PE positive population, neutrophil specific CD66b-APC-Alexa Fluor 750 positive population, monocytes specific CD14-PE positive population and lymphocytes specific CD3-ECD positive population. A number of 20,000 events were acquired and analyzed using CytExpert software to determine the percent and mean fluorescence intensities (MFI) of analyzed cell subsets.

### Measurement of ROS by flow cytometry

The intracellular ROS generation by different cell populations was measured in whole blood samples using 2’,7’-dichlorofluorescein-diacetate (DCF, Sigma Aldrich, D6883). Suspended cells were incubated with DCF (20 μM) and combinations of monoclonal antibodies; CD-42b-PE (Platelets), CD14-PC7 (monocytes), CD66b-APC-Alexa Fluor 750 (neutrophils), and CD3-ECD (lymphocytes) for 30 minutes at room temperature in the dark. Cells were then washed with PBS and resuspended in 300 μl of PBS. 20,000 events were recorded and analyzed using CytExpert program.

### Determination of hydrogen peroxide concentrations in plasma

Catalase was used to specifically and quantitatively determine levels of hydrogen peroxide in identical plasma volumes collected from the study subjects. Oxygen levels are monitored and recorded while 50 μL batches of plasma from control, Sev-R, and Sev-D subjects sequentially infused into tightly air-controlled O2k chamber containing catalase (315 Units/mL) in deoxygenated buffer. A continuous stream of pure nitrogen gas was blown over the chamber’s sealing cap to prevent oxygen diffusion into the working solution. In addition to the initial rise due to residual oxygen in the added plasma samples, the decomposition of hydrogen peroxide in these samples produced oxygen in a quantitative manner; i.e. decomposition of one mole of H_2_O_2_ produced ½-mole O_2_. To verify the assay, we measured the released oxygen upon adding increasing concentrations of standard hydrogen peroxide solution in PBS buffer. Linear fitting of the plotted [O_2_] versus [H_2_O_2_] relation yielded a slope = 0.47 ± 0.03 (Pearson’s r = 0.994, *p* = 5.6 × 10^−4^), which is very close to the theoretically expected value of 0.5.

### Measurement of ROS in isolated neutrophils by fluorescence imaging

The intracellular ROS generation of isolated neutrophils was detected and quantified using the ROS-sensitive DCF dye. Isolated cells were seeded and incubated for 30 minutes at 37°C followed by centrifugation to form a monolayer. Cells were then stained with DCF at a final concentration of 0.3 mM in PBS for 30 minutes. Cells were washed and stained with DAPI (Hoechst 33342 Solution, ThermoFischer Scientific, 62249) at a final concentration of 10 μM in PBS for 30 minutes at 37°C. Cytation 5 Cell Imaging Multi-Mode Reader (Agilent) was used to acquire images using 20X lens and the proper fluorescence filter cubes (λ_ex_ = 500 ± 12 nm and λ_em_ = 542 ± 14 nm). Images were processed to quantify fluorescence intensity using Gen5 software package 3.08.

### Determination of plasma albumin concentrations

Albumin concentration in plasma was measured using bromocresol green (BCG) dye as the color intensity of the complex formed is proportional to albumin concentration. Standard curve was generated using standard albumin and samples were measured in 96 well plate at 630 nm using Cytation 5 (Agilent). Concentrations were calculated using the equation generated from standard curve and presented in mg/mL.

### Statistical analysis

Statistical analysis and data graphing were performed using OriginPro 2017 (OriginLab Corporation, Northampton, USA). For comparisons of means between three or more independent groups, Tukey test ANOVA for multiple comparison was performed. For correlation analysis, spearman rank correlation was performed. Pearson’s correlation coefficient was calculated for measuring the association between variables of interest based on the method of covariance which also gives information about the magnitude of the association, or correlation, as well as the direction of the relationship. *p*-values < 0.05 for correlations or means’ comparisons were considered significant. Categorical variables are reported as counts and percentages while continuous variables are expressed as mean ± standard deviation. Differences between percentages were assessed by Pearson’s χ^2^ tests or Fisher exact tests when the number of observations per group were less than 5. The χ^2^ tests provided results that tested the hypothesis that the mortality and a given variable (e.g. sex or a comorbidity) are independent. When *p* is less than the significant level of 0.05, there is significant evidence of association between mortality and the variable. Log-rank Kaplan–Meier survival analyses were carried out to estimate probability of survival of COVID-19 patients in relation with cut-off thresholds arbitrarily stratified at mean values of various parameters. The log-rank test for trends reports a χ^2^-value and computes a *p* value testing the null hypothesis that there is no linear trend between column order and median survival.

### Data sharing statement

Original datasets and detailed protocols are available upon request from the corresponding author.

## Results

### Demographic, clinical, and laboratory hematologic characteristics of COVID-19 patients

**Table 1** lists demographic data, comorbidities, ongoing medications, and administered anti-COVID-19 medications applied to treat current study participants that were divided into survivors (Sev-R) and deceased (Sev-D). Compared with recovered, deceased patients’ mean age was slightly older (Tukey ANOVA, *p*<0.05). No other clinical or demographic characteristic showed statistically significant difference between Sev-R and Sev-D groups when analyzed by Pearson’s Chi-Square test except that the frequency of remdesivir administration was significantly higher in Sev-D groups (p<0.05). In **Table 2** we show and statistically compare laboratory results of survivors vs. non-survivor COVID-19 groups. Although when comparing all parameters in the two COVID-19 groups we observed changes following the same reported trends in the literature, means’ comparisons by Tukey test reported non-significant changes with strong trends observed only for C reactive protein (greater levels in Sev-D group, *p* = 0.050) and albumin (lower levels in Sev-D group, *p* = 0.067). Nevertheless, non-survivors’ blood carried the frequently observed hallmarks of increased CRP, D-dimer, IL-6, ferritin, and the liver enzymes ALT and AST (Reviewed in: (27, 28)). However, it is concievable that the clinical severe category and the same ICU status of patients in the two groups in addition to relatively small sample size underlie the observed lack of robust statistical differences between these parameters.

**Table 2.** Laboratory parameters of the current study patients.

|  | **Sev-R**<br>**(mean ± SD)** | **Sev-D**<br>**(mean ± SD)** | **Tukey 95%-CI** | ***p*** |
| --- | --- | --- | --- | --- |
| **WBCs (x 10<sup>3</sup>/mL)** | 10.85 ± 4.3 | 12.5 ± 5.1 | -2.62–5.94 | 0.42 |
| **Platelets (x 10<sup>6</sup>/mL)** | 274.3 ± 88.3 | 233.1 ± 92.2 | -116.7–34.1 | 0.27 |
| **INR** | 1.34 ± 0.63 | 1.26 ± 0.23 | -0.49–0.33 | 0.68 |
| **CRP (mg/L)** | 39.95 ± 42.6 | 90.6 ± 66.9 | -0.1–101.4 | 0.05 |
| **D-dimer (µg/mL)** | 1.38 ± 1.7 | 3.79 ± 4.6 | -0.81–5.64 | 0.13 |
| **IL-6 (pg/mL)** | 214.8 ± 442.6 | 393.0 ± 749.6 | -427–783 | 0.54 |
| **Ferritin** | 715.6 ± 426 | 1081 ± 622 | -94–826 | 0.11 |
| **Albumin (g/mL)** | 31.8 ± 4.8 | 26.9 ± 5.1 | -1.0–0.04 | 0.067 |
| **Hemoglobin (g/dL)** | 12.18 ± 1.3 | 12.16 ± 2.0 | -1.5–1.46 | 0.97 |
| **ALT (U/L)** | 29.6 ± 20.2 | 40.1 ± 37.3 | -15.4–36.4 | 0.40 |
| **AST (U/L)** | 34.7 ± 20.5 | 44.3 ± 29.8 | -12.2–31.4 | 0.37 |
| **Serum creatinine**<br>**(mg/dL)** | 1.24 ± 0.8 | 1.78 ± 1.7 | -0.64–1.73 | 0.35 |

WBCs, white blood cells; INR, international normalized ratio; CRP, high-sensitivity C reactive protein; ICU, intensive care unit; PLTs, platelets; ALT, alanine transaminase; AST, aspartate transaminase. The Tukey's calculated *p*-values as well as upper and lower 95%-confidence levels for the Sev-R vs. Sev-D means' comparisons are given.

### Neutrophils are a major source of reactive oxygen species

It has been recently proposed that the high neutrophil-to-lymphocyte ratio observed in critically ill COVID-19 patients may tip the redox homeostasis due to increased reactive oxygen species production (15). Our hypothesis implicates elevated oxidative stress as a major cause of HSA damage in severe COVID-19 patients. As a result, we started by following the dependence of clinical outcomes and mortality on ROS levels in blood cells. First, we used flowcytometry to assess percentages of neutrophils, lymphocytes, and platelets in all patients as described in Materials and Methods (Fig. 2A). Furthermore, we used the ROS-sensitive DCF dye to probe intracellular ROS levels in various cell populations in whole blood from all groups. Fig. 2 shows that while lympocyte counts decrease, a parallel dramatic increase in neutrophil counts (% total) was observable when going from Control (40.78 ± 14.0, n=9) to Sev-R (64.0 ± 20.0, n=10) to Sev-D (76.4 ± 6.8, n=11) groups (overall ANOVA *p* = 3.9 × 10^−5^). Similar trend was clearly seen in the heat map depicting parameters for all patients analyzed by flow cytometry (Fig. 2B). It is also clear from Fig. 2B&C that changes in DCF-positive neutrophils follows similar trend observed for neutrophil counts. To confirm this relation we compared neutrophil counts with DCF-positive neutrophil counts and found that the two parameters were strongly correlated (Pearson’s r = 0.8, *p* = 3 × 10^−7^), Fig. 2C. Moreover, both parameters individually showed statistically significant increases in both of the studied COVID-19 groups when compared with control group (Fig. 2D&E). These results suggest that neutrophils are major sources of elevated oxidative stress in critically ill patients. Note that the observed trends in platelets, lymphocyte, neutrophils, and neutrophil-to-lymphocyte ratio (NLR) are similar to reported values (29, 30).

**Fig. 2.**
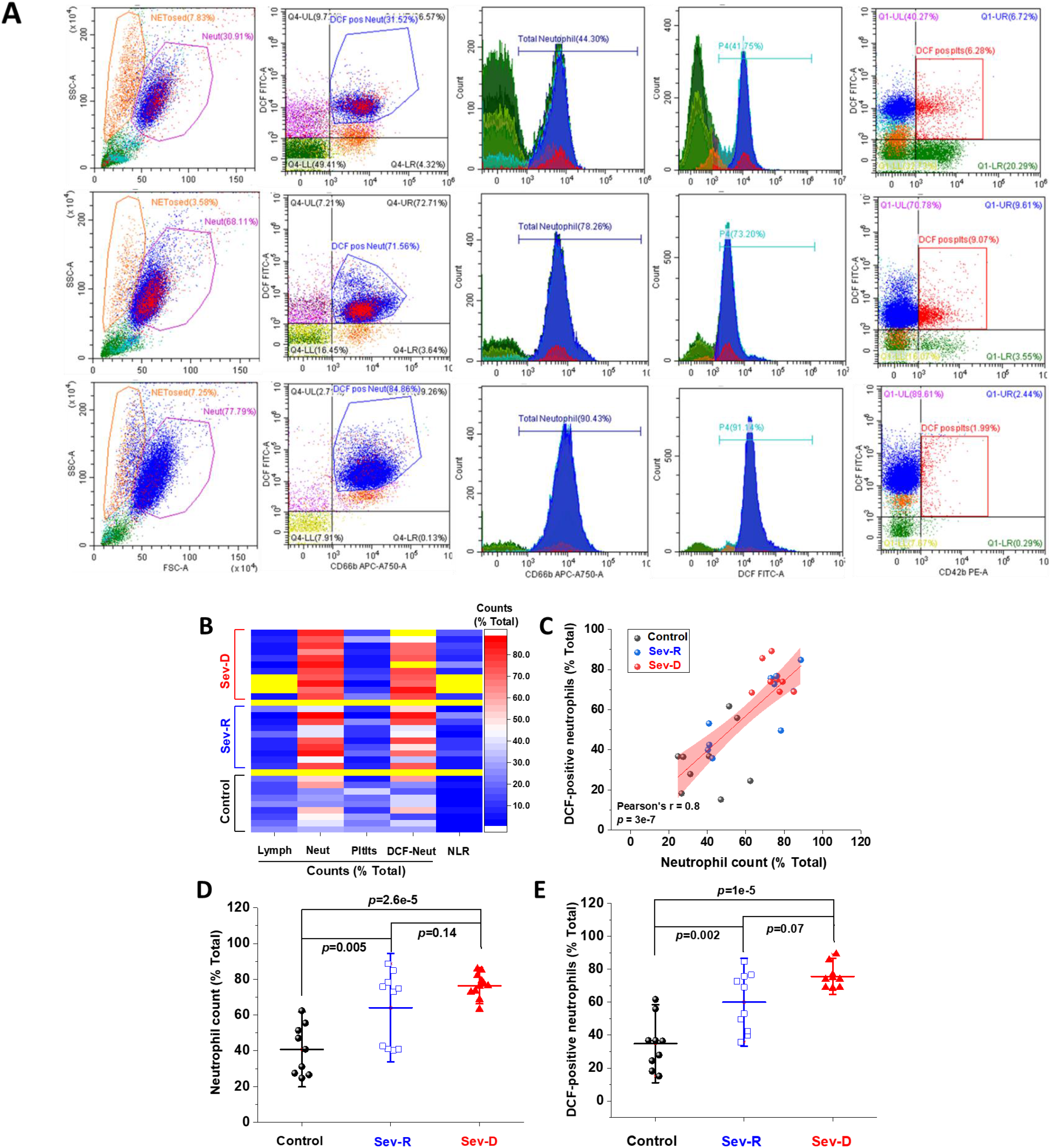
Hematologic cellular counts and neutrophil-ROS levels reflect severity and mortality in COVID-19 patients. A) Representative flow cytometric diagrams comparing morphologic, hematologic, and ROS levels in control (representative of n=9; upper row), Sev-R (representative of n=10; middle row), and Sev-D (representative of n=11; lower row) groups. B) Heat diagram comparing lymphocyte, neutrophils, platelets, and DCF-positive neutrophil counts as percentage of total cell counts in all of the studied subjects. Yellow areas are either group separators or missing data due to insufficient sample size or processing errors. C) A diagram showing statistically positive correlation between neutrophil count and count of neutrophils stained positive for DCF dye in all groups (black dots denote controls; blue are Sev-R; and red represent Sev-D patients). D) When neutrophil counts were compared for all groups, both Sev-R and Sev-D groups showed statistically significant neutrophilia relative to control groups. However, only a weak trend has been observed when comparing the two groups with COVID-19. E) DCF staining revealed increased levels of ROS in Sev-R and Sev-D groups relative to control neutrophils. Sev-D showed a trend of increased ROS level relative to Sev-R group. Multiple comparisons were carried out using ANOVA followed by Tukey test and *p* values are given.

### Hydrogen peroxide levels in plasma correlate with mortality

Next, we reasoned that elevated oxidative stress in both groups with critical COVID-19 infection would be echoed in plasma levels of hydrogen peroxide. We used a highly specific catalase-based assay that we developed and verified in our laboratory to quantify [H_2_O_2_] in plasma samples of all groups. The assay relies on high resolution detection and quantification of released oxygen due to hydrogen peroxide decompostion by catalase, Fig. 3A&B. We constructed a calibration curve to confirm the catalase-mediated H_2_O_2_ to O_2_ stoicheometric conversion (Fig. 3B). A linear relation was obtained with zero intercept and slope of 0.47±0.03 which closely matches the theoretically expected value of 0.5 (95% CI: 0.37-0.56, *p* = 5.6 × 10^−4^, Pearson’s *r* = 0.994). Indeed, we detected striking differences between groups even with relatively small sample sizes (Mean±SD, Control, n=7: 2.57±0.57, Sev-R, n=6: 5.37±1.0, Sev-D, n=7: 8.8±1.7 ; overall ANOVA *p*-value = 1.15 × 10^−7^ ; Fig. 3C). The differences between groups have reached statistical significance (Sev-R vs. Control, 95% CI: 1.08-4.51, *p* = 0.0017; Sev-D vs. Control, 95% CI: 4.58-7.87, *p* = 0.0; Sev-D vs. Sev-R, 95% CI: 1.72-5.15, *p* = 2.3 × 10^−4^). It appears from these results that a measure of oxidative stress; *i.e*. [H_2_O_2_] in plasma is doubled in survivors and quadropled in deceased COVID-19 patients relative to controls’ plasma average levals.

**Fig. 3.**
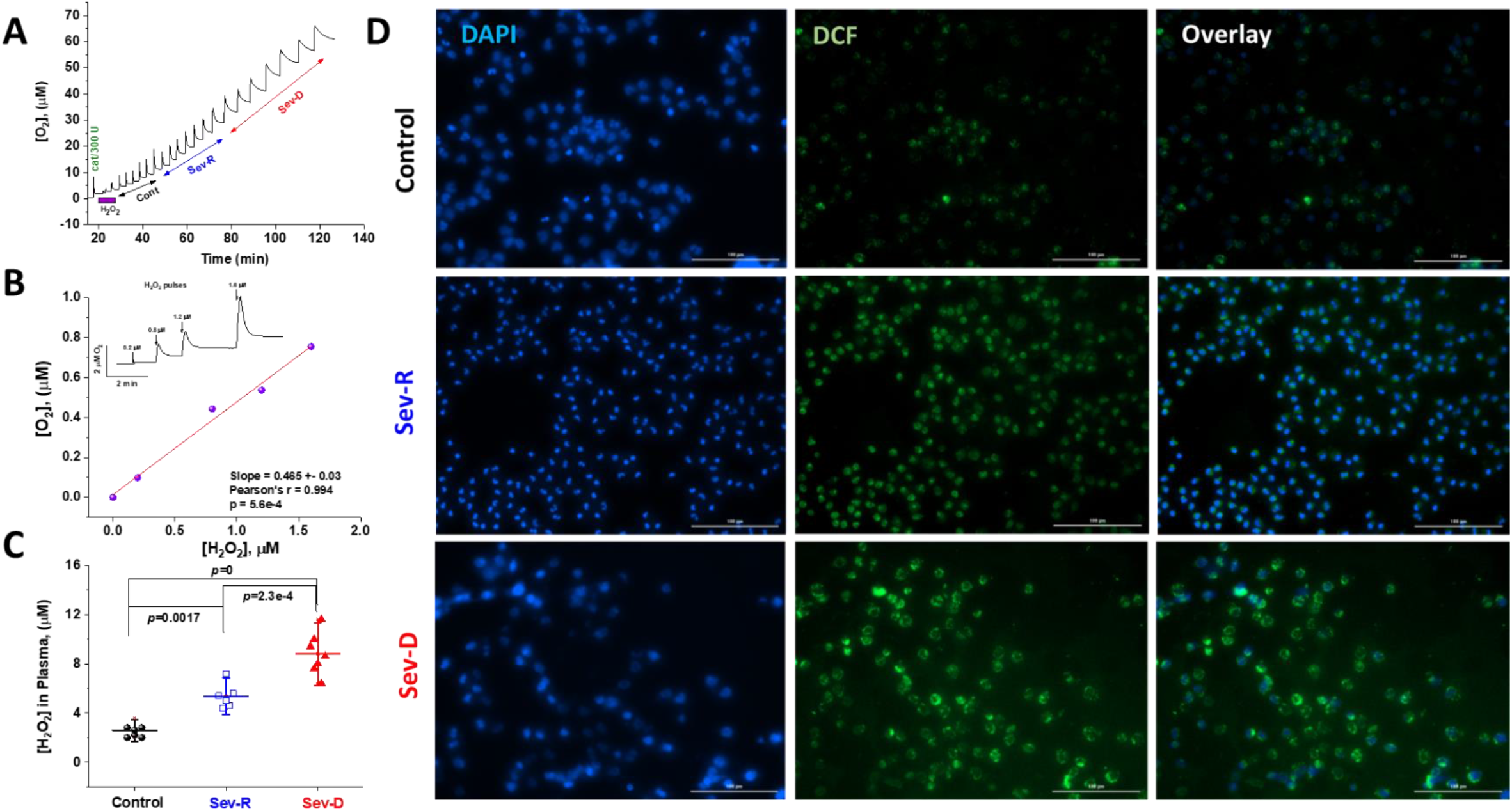
Hydrogen peroxide levels in plasma and neutrophils reflect mortality in COVID-19 patients. Catalase was used to specifically and quantitatively determine levels of hydrogen peroxide in identical plasma volumes collected from control (n=7), Sev-R (n=6), and Sev-D (n=7) groups. A) Oxygen levels are monitored and recorded while 50 mL batches of plasma from control, Sev-R, and Sev-D subjects sequentially infused into tightly air-controlled O2k chamber containing catalase (315 Units/mL) in deoxygenated buffer. In addition to the initial rise due to residual oxygen in the added plasma samples, the decomposition of hydrogen peroxide in these samples produces oxygen quantitatively. B) To verify the assay we measured the released oxygen upon adding an increasing volume of standard hydrogen peroxide solution in PBS buffer with 0.2, 0.8, 1.2, and 1.6 μM final concentrations; inset. Linear fitting of the plotted [O_2_] versus [H_2_O_2_] relation yielded a slope = 0.47 ± 0.03 (Pearson’s r = 0.994, p = 5.6 × 10^−4^), which is very close to the theoretically expected value of 0.5 as the catalase-mediated decomposition of one mole of H_2_O_2_ produces ½-mole O_2_. C) Plasma contents of H_2_O_2_ in plasma significantly increased in the order Sev-D>Sev-R>Cont using ANOVA followed by Tukey test. D) Fluorescence imaging was used to assess levels of ROS in freshly isolated neutrophils using DCF (2,7-Dichlorodihydrofluorescein diacetate, green) staining in all groups. DAPI (4′,6-diamidino-2-phenylindole, blue) binds strongly to adenine– thymine-rich regions in DNA thus mapping nuclei through emitting blue fluorescence. Merged DCF and DAPI images are shown on the third column. Images were acquired using Cytation 5 Cell Imaging Multi-Mode Reader (Agilent) and analyzed using Gen5 Software package 3.08. Scale bar: 100 µm.

To confirm this finding we performed DCF fluorescence imaging on freshly isolated neutrophils of representative group of individuals from each group, Fig. 3D. We simultaneously stained neutrophils’ nuclei with DAPI (blue stain) to follow nuclear morphologic changes and DNA diffusion in all groups. Although requiring more detailed studies, a closer look on the acquired images of DAPI-stained neutrophils from a survivor patient showed significantly reduced average neutrophil size (control DNA area, 144.8 ± 93.7 μm^2^ (133 cells analyzed) vs. Sev-R DNA area, 36.0 ± 7.5 μm^2^, (241 cells analyzed), Welsh corrected two samples t test *p* = 0) with tendencies towards more condensed, more segmented nuclei and frequent C shaped chromatin. However, neutrophils from a non-survivor exhibited diffused chromatin and possessed in average a 26% larger DNA area than normal cells and roughly 5 times that of Sev-R neutrophils (DNA area (97 cells analyzed), 182.7 ± 171 μm^2^, *p* = 0 *vs*. both control and Sev-R groups). Inspection of the DCF flourescence images indicated that the non-survivor’s neutrophils contained larger populations of what appears to be toxic granules and cytoplasmic vacuoles that are highly ROS-positive. Analysis of mean DCF fluorescence intensities (MFI) per cell confirmed results obtained by flow cytometry and catalase assay reporting increased levels of ROS in the order control << Sev-R < Sev-D (DCF MFI ± SD (x 10^3^): Control (282 cells analyzed), 6.7 ± 0.7; Sev-R (351 cells analyzed), 10.9 ± 0.9; Sev-D (368 cells analyzed), 13.0 ± 1.1, Welsh corrected two samples t test, *p* = 0 for all comparisons).

### EPR-determined biophysical parameters reflecting albumin conformational changes are consistent predictors of COVID-19 mortality

It has been previously shown that non-survivor COVID-19 patients exhibit mild but consistent hypoalbuminemia relative to survivors (31). We started by assessing albumin levels in the studied cohort of subjects to confirm if they follow similar trends. We found that [albumin] in plasma decreased in the order Control > Sev-R >> Sev-D (Mean±SD, Control, n=8: 40.45±10.93 mg/mL, Sev-R, n=8: 30.66±10.99 mg/mL, Sev-D, n=10: 24.71±5.88 mg/mL; overall ANOVA *p*-value 0.006; Fig. 4A). We only detected statistically significant decrease in [albumin] in plasma of Sev-D group (Sev-R vs. Control, 95% CI: −21.44-1.87, *p* = 0.11; Sev-D vs. Control, 95% CI: −26.79--4.67, *p* = 0.004; Sev-D vs. Sev-D, 95% CI: - 17.0-5.11, *p* = 0.38). The reference range of HSA concentration in serum is approximately 35-50 mg/mL, but we and other groups found that COVID-19 associated mortality correlates with lowered [albumin] < 30 mg/mL (31). However, high prevelance of hypoalbuminemia in numerous disease states and the age/sex-dependent wide dynamic range of this protein concentration limits its diagnostic utility (32). As a result, we investigated biophysical parameters pertaining to HSA protein configuration in whole blood and plasma of all groups as reflectors of this critical protein functions.

**Fig. 4.**
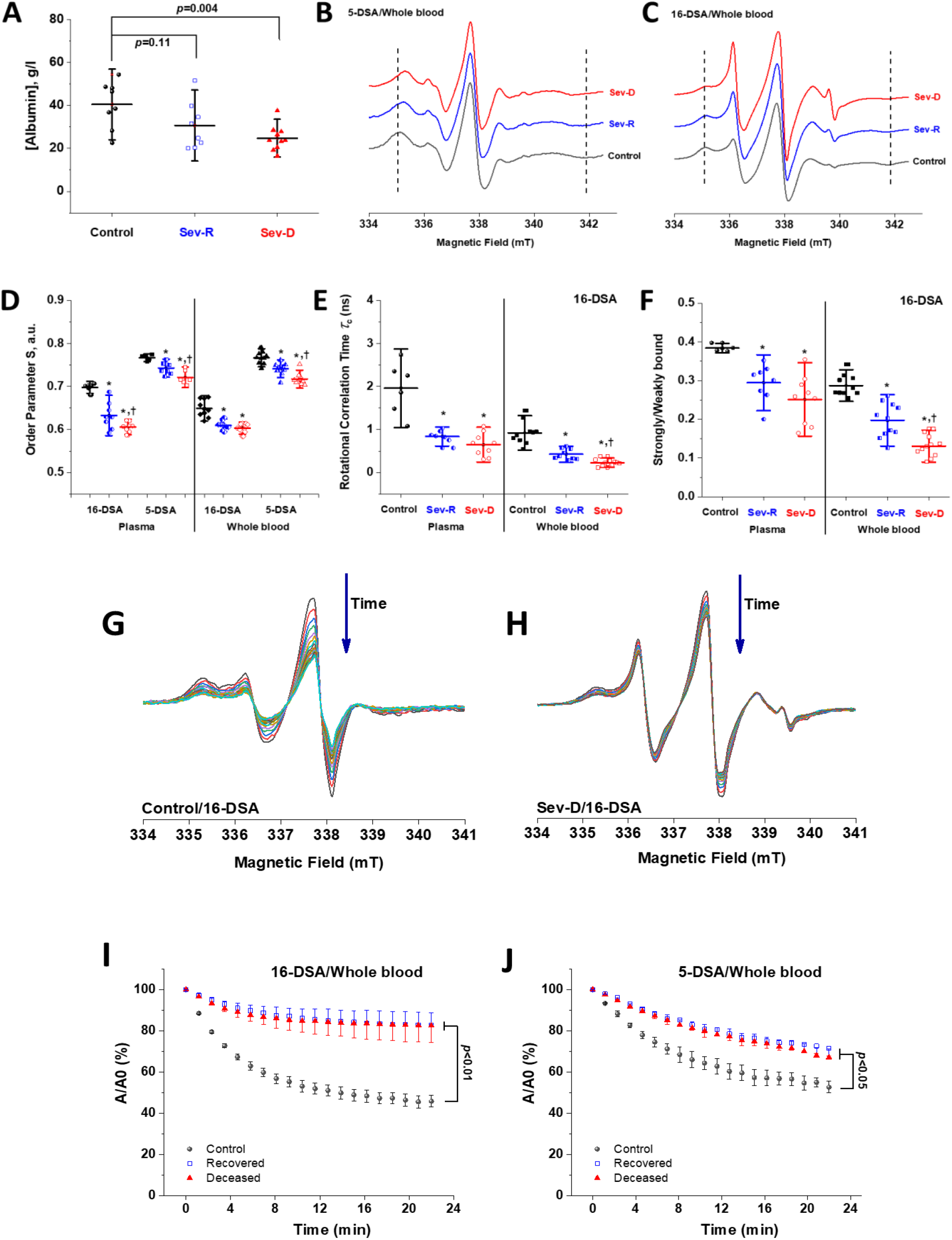
EPR spectroscopic analyses of HSA-fatty acid binding reveal strong dependence of binding on mortality in COVID-19 patients. A) Albumin level in plasma of control (n=8), Sev-R (n=8), and Sev-D (n=10) groups showed that nonsurvivor COVID-19 patients exhibit statistically significant hypoalbuminemia. Comparisons between representative spectra showing changes in line shape that are related to mobility and microenvironmental statuses of HSA-bound 5-DSA (B) and 16-DSA (C) in whole blood of a control (black trace), a Sev-R (blue trace), and a Sev-D (red trace) patients. Calculated biophysical parameters including order parameter (D), rotational correlation time (E), and the ratio between strongly bound to weakly bound spin labels (S/W, F) as defined in Fig. 1 and described in Methods section. Statistical comparisons by ANOVA followed by Tukey tests were used for means’ comparisons and revealed remarkable decrease in the binding strengths and packing parameter of the local microenvironment surrounding the spin labels. All calculated parameters along with exact *p* values are given in the Supplementary Table 3. Water accessibility into albumin/fatty acids binding pockets are followed by reacting with ascorbate, which reduces nitroxide radicals into the EPR silent hydroxylamines. Representative EPR signal decays of 16-DSA in whole blood of control (G) and Sev-D (H) are shown. Kinetic traces (n=3 per group) showing the reduction of 16-DSA (I) and 5-DSA (J) bound to HSA by sodium ascorbate in whole blood. Kinetic traces are shown as the percentage loss of the signal intensity of the middle peak (A/A_0_). All samples contained 0.26 mM spin label and 3 mM sodium ascorbate and measured at 37 °C. Weaker and slower disappearance of the EPR signal suggests inaccessible space towards the nitroxide moiety of the spin label.

Previous studies reported that allosteric changes in HSA may be utilized to reflect critical functional changes in albumin and explored diagnostic and prognostic values of these changes in cancer (17, 33). It has also been found that long chain fatty acids binding alters the interactive binding of ligands in the two major drug binding sites of HSA (34). We employed EPR spectroscopy to probe SLFAs’ binding statuses and protein configuration in all groups as detailed in the Methods’ section above. Analysis of the 5-DSA and 16-DSA EPR spectra revealed remarkable changes in spectral features between control and COVID-19 groups (Fig. 4B, 5-DSA; Fig. 4C, 16-DSA in whole blood). For example, the hyperfine coupling tensor element 2*T*_‖_ (defined in Fig. 1B and used to calculate the protein packing order parameter *S*) is sensitive to microenvironmental effects (such as polarity, H-bonding, electrostatic interactions, etc.) on the spin probe that is localized in one of the HSA native fatty acids binding pockets. Also, the S/W ratio (see Fig. 1C) corresponding to strongly-bound/weakly-bound populations of 16-DSA spin probe may reflect changes in protein folding that can alter protein-fatty acid interactions. Furthermore, the rotational correlation time *τ*_c_ which is a measure of the spin probe rotational mobility is also analyzed.

Calculated EPR spectral parameters for all groups are listed in Supplementary Table 1 including exact ANOVA and Tukey test *p* values along with the number of subjects analyzed. Both 5-DSA and 16-DSA were used to probe the degree of local interactions in sites where the spin labeled fatty acid is buried in the protein interior (high mobility, 16-DSA) and closer to the protein-aqueous interface (low mobility due to interactions with water molecules and polar amino acids, 5-DSA)(35). Indeed, 5-DSA reflected significantly greater *S* parameter values relative to 16-DSA both in plasma and in whole blood (Fig. 4D). However, independent of the spin probe and both in plasma and whole blood samples, the order parameter has been consistently lower in Sev-R which was further decreased in Sev-D patients relative to control group (Fig. 4D). In whole blood, similar results that showed more statistically robust differences have been observed.

Calculations of *τ*_c_ as described in Methods showed rotational mobility of 16-DSA is significantly faster when bound with HSA from COVID-19 patients relative to that from control subjects in plasma or whole blood. Finally, similar trends have been observed for the S/W parameter which signifies contributions of the strongly and weakly bound components of 16-DSA in different fatty acids pockets. Taken together, these results indicate that COVID-19 pathology is associated with extensive structural changes in the HSA protein that imply the prevelance of malfunctional derivatives of this critical protein.

### Hampered water accessibility into HSA/fatty acid pockets in whole blood of COVID-19 patients

We followed water accessibility towards deep pockets carrying the spin labels through kinetic analysis of the nitroxide radical EPR silencing by the water-soluble ascorbate anion (36); Fig. 4(G-J). Under matching experimental conditions, 16-DSA and 5-DSA/HSA signals in whole blood of control subjects decayed remarkably faster when compared with both Sev-R and Sev-D (n=3 per group, p<0.05). Weaker and slower disappearance of the EPR signal of COVID-19 patients by ascorbate suggests less accessible space towards the nitroxide moiety of the spin label within the protein. It is clear from these data that HSA of COVID-19 patients is generally less water-accessible relative to control ones. However, the core of the HSA of both COVID-19 groups were not significantly different in terms of water-accessibility.

### Association of EPR-determined HSA structural changes with neutrophil counts and measures of oxidative stress

In order to verify our proposed links between oxidative stress and albumin damage we applied linear correlations between plasma levels of hydrogen peroxide and EPR-calculated parameters pertaining to protein packing order parameter (S), fatty acid mobility (*τ*_c_), and S/W ratio in all groups (n=19-26, Fig. 5A-D). Furthermore, to substantiate our suggestion that neutrophils are major sources of oxidative stress implicated in COVID-19 severity and mortality, we explored linear correlations between the populations of DCF positive neutrophils and *τ*_c_ (Fig. 5E) or S/W ratio (Fig. 5F). Despite relatively small numbers of subjects analyzed, correlations of biophysical parameters showed moderate-to-strong negative correlations with [H_2_O_2_] in plasma or with DCF positive populations (% of total); Pearson’s r = −0.6- −0.76, p < 0.01 for all correlations. These observations may suggest that neutrophil-mediated oxidative stress in critically ill COVID-19 patients is a potential contributor to the observed structural changes in HSA of those patients.

**Fig. 5.**
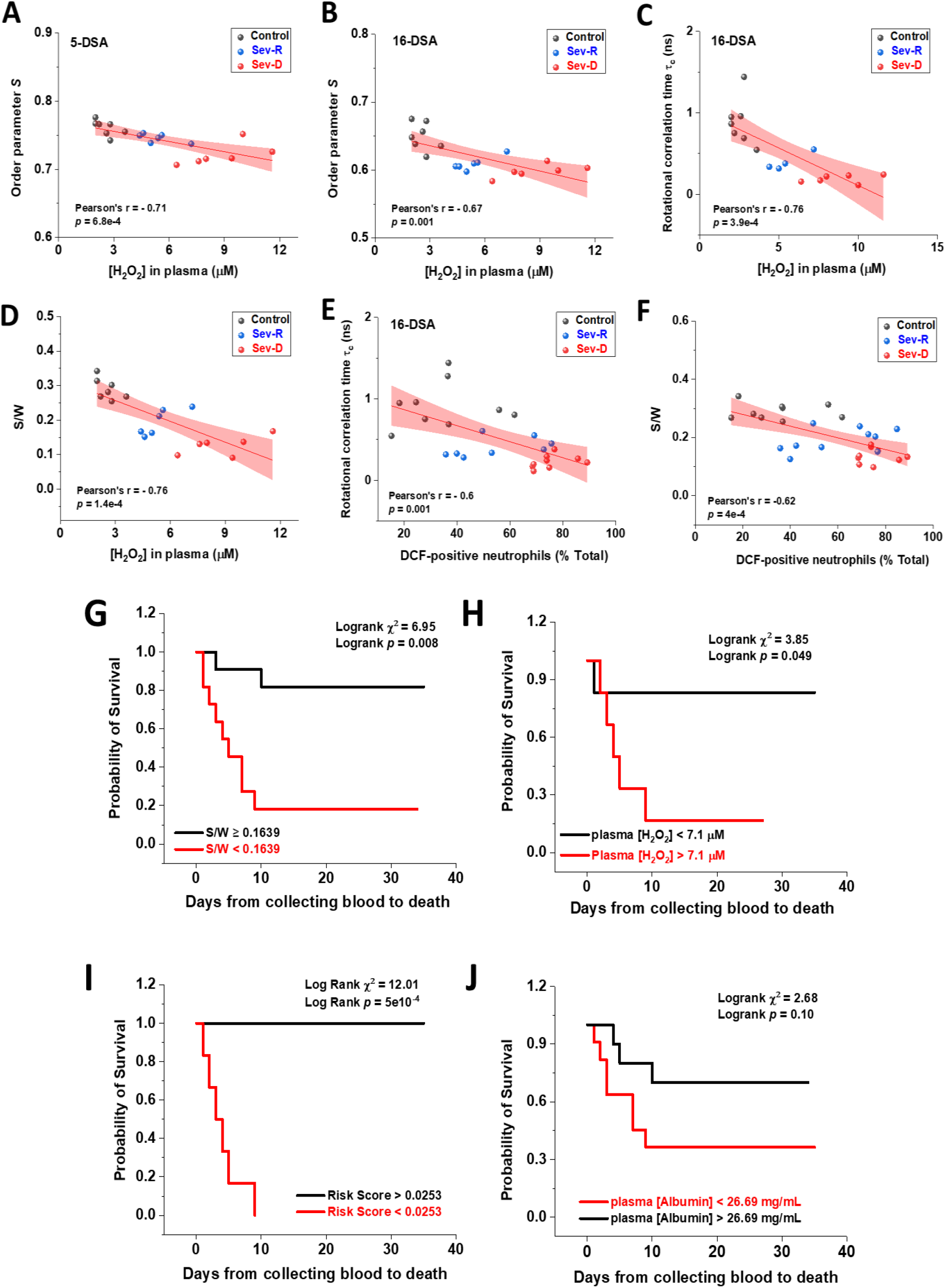
Associations of oxidative stress measures and biophysical parameters with mortality outcome in COVID-19 subjects. Linear correlations between plasma levels of hydrogen peroxide and EPR-calculated parameters pertaining to protein packing order parameter (A,B), fatty acid mobility *τ*_c_ (C), and S/W ratio (D) in Control (black circles), Sev-R (blue circles), and Sev-D (red circles) groups (n=19-26). Neutrophils are major sources of oxidative stress as evident by linear correlations between % DCF positive neutrophils and *τ*_c_ (E) or S/W ratio (F). On each correlation, Pearson’s r and *p* values are provided. **Kaplan–Meier estimates of time to mortality from blood sample collection during ICU hospitalization (G-J)**. Logrank Kaplan–Meier survival analyses were carried out to estimate probability of survival of COVID-19 patients in relation with cut-off thresholds arbitrarily selected as the mean values of the analyzed parameters. For S/W, plasma [H_2_O_2_], Risk Score defined as the {(S/W)/[H_2_O_2_]} ratio, and plasma [Albumin], the number of analyzed COVID-19 patients were 22, 12, 12, and 21; respectively.

### Analysis of Kaplan–Meier estimates of time-to-mortality from blood sample collections during ICU hospitalization

Finally, we stratified patients into two groups, patients showing values below the means and patients with values above the means of each of the studied parameters: (*i.e*. S/W mean=0.1639 (0.090-0.261); plasma [albumin] mean = 0.26.69 mg/mL (16.42-51.48); and plasma [H_2_O_2_] mean = 7.1 μM (4.4-11.6)). Patients with lower values of the S/W ratio showed significantly higher in-hospital mortality (81.8% vs. 18.2%, logrank χ^2^ = 6.95, p = 0.008, Fig. 5G). Similarly, patients with accumulated H_2_O_2_ in their plasma showed higher mortality (83.3% vs. 16.7%, logrank χ^2^ = 3.85, p = 0.049, Fig. 5H). However, when we combined these two parameters to derive a risk score as the ratio ((S/W)/[H_2_O_2_]), the resultant risk score lower than the mean (< 0.0253) predicted mortality with 100% accuracy (100% vs. 0%, logrank χ^2^ = 12.01, p = 5×10^−4^, Fig. 5I). This consistent statistics must be verified over larger sample size which is currently in pursuit. Under our current conditions, although showing weak statistical trends both of the order parameter (not shown) and the plasma albumin level (Fig. 5J) were not statistically significant determinants of in-hospital mortality.

## Discussion

Human serum albumin is a pivotal protein with diverse multifaceted functions that are increasingly reported to reflect physiological and contribute to pathological states (32, 37). Closer to the context of COVID-19 pathophysiology, critical roles of HSA have been suggested as it acts as an anti-inflammatory and antioxidant protein (38), anticoagulant agent (39, 40), inhibitor of oxidative stress-mediated clotting and platelet activation (41, 42), drug-dependent allosteric carrier and regulator (43), and a potent heme scavenger that may also exhibit globin-like reactivity (44). In the light of these established functions it is natural to suggest albumin as a frontline protection against COVID-19-associated lethality resulting from cytokine storm, oxidative stress, blood clotting, and the ensuing organ failure. Although hypoalbuminemia has been repeatidly reported as a predictor of mortality in COVID-19 (31, 45), low albumin levels may result from surgery, dialysis, abdominal infections, liver failure, pancreatitis, respiratory distress, bypass surgery, ovarian problems caused by fertility drugs, and many other conditions. High prevelance of hypoalbuminemia in numerous disease states and the age/sex-dependent wide dynamic range of this protein concentration limits its diagnostic utility (32). As a result, we investigated biophysical parameters pertaining to HSA protein structure in whole blood and plasma of all groups as reflectors of this critical protein functions.

We designed the present study to explore how COVID-19-caused mortality is related to oxidative stress and whether ROS-induced albumin damage can be used to predict such outcome. To avoid disparities related to clinical statuses and interventional protocols we restricted our subject recruitment to a homogeneous cohort of patients in terms of diseases severity, hospitalization, and interventions. Both sexes were represented in the overal COVID-19 subjects (44% females) and sex was not found to affect mortality (*p* = 0.86). The mean age of the studied subjects was 66.7 ± 8.9 y (42-81), and non-survivors were slightly orlder (p = 0.04). No other clinical or demographic characteristic showed statistically significant difference between survivors and non-survivors which further indicates the homogeniety of the studied pool of subjects.

Our current data provide converging and novel evidence that neutrophils are major players in oxidative stress associating inflammation in critically ill COVID-19 patients, especially those that don’t survive the infection. We detected increased neutrophil count, increased ROS-positive neutrophil population, increased intracellular ROS level in these neutrophils, and a remarkably elevated hydrogen peroxide residue in plasma of non-survivor COVID-19 patients relative to control and survivor groups. Moreover, these oxidative stress measures were found to strongly correlate with EPR-detected strucrtural damages of HSA. Although we found a weak trend that albumin levels were inversely associated with overall mortality, a biophysical measure of structural changes inflicted on albumin (S/W) was found to consistently predict overall mortality (*p* = 0.008) even with a relatively small n=22 sample size. Moreover, plasma levels of hydrogen peroxide was also found to predict in-ICU mortality with even smaller number of analyzed samples (n=12, *p*=0.049). This substantiates the utilization of these parameters as accurate mortality predictors of critically ill COVID-19 patients. Determined threshold values of these parameters can possibly help identifying patients in need for urgent medical attention, and may provide novel markers to assess new candidates for COVID-19 treatments targeting HSA replacements. We thus suggest the importance of studying the potential effects of albumin replacement therapy on clinical outcomes of Covid-19 patients. Our results also provide new mechanistic insights into pathways that could be targeted to help prevent critical COVID illness and death; e.g. oxidative stress.

## Data Availability

Original datasets and detailed protocols are available upon request from the corresponding author.

## Acknowledgments

The present work was funded by the Association of Friends of the National Cancer Institute and the Children’s Cancer Hospital Foundation.

## Authorship Contributions

MAB, BAY, EAA-R, AAE, and AK performed experiments, analyzed results, and assisted in the manuscript preparation. Subjects’ recruitment, ethical approvals, clinical follow-up and assessments were coordinated by RE-M. AMS, RH, DAl-R, MZ, and NE assisted in experimental work. MH, MEl-A, AEl-H, AE, AH, LLD and SA provided logistical support and critical manuscript revisions. SSA conceived and directed the project, designed experiments, analyzed data, and wrote the manuscript.

