## Supplementary figures and images for "Neutrophil-mediated Oxidative Stress and Albumin Structural Damage Predict COVID-19-associated Mortality" (from another version of this preprint: version 1, 2021-04-07)

### D:\Sameh Ali-D drive\57357 papers\COVID papers\Albumin\For Submission\JCI

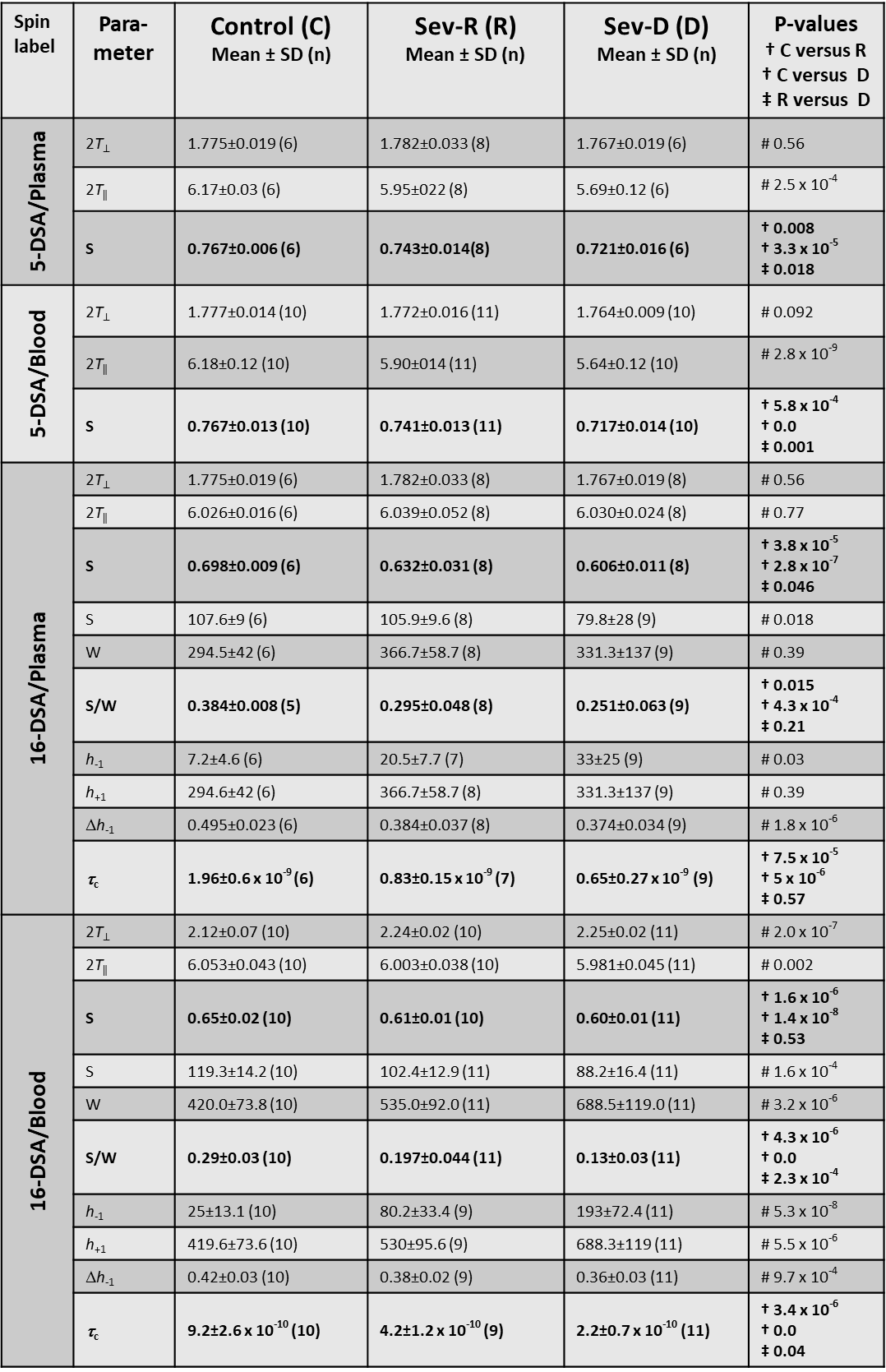

**Supplementary Table 1. Calculated biophysical EPR spectral parameters for all groups .**
